# Enhancing Dietary Supplement Question Answer via Retrieval-Augmented Generation (RAG) with LLM

**DOI:** 10.1101/2024.09.11.24313513

**Authors:** Yu Hou, Rui Zhang

## Abstract

**Objective:** To enhance the accuracy and reliability of dietary supplement (DS) question answering by integrating a novel Retrieval-Augmented Generation (RAG) LLM system with an updated and integrated DS knowledge base and providing a user-friendly interface. With.

**Materials and Methods:** We developed iDISK2.0 by integrating updated data from multiple trusted sources, including NMCD, MSKCC, DSLD, and NHPD, and applied advanced integration strategies to reduce noise. We then applied the iDISK2.0 with a RAG system, leveraging the strengths of large language models (LLMs) and a biomedical knowledge graph (BKG) to address the hallucination issues inherent in standalone LLMs. The system enhances answer generation by using LLMs (GPT-4.0) to retrieve contextually relevant subgraphs from the BKG based on identified entities in the query. A user-friendly interface was built to facilitate easy access to DS knowledge through conversational text inputs.

**Results:** The iDISK2.0 encompasses 174,317 entities across seven types, six types of relationships, and 471,063 attributes. The iDISK2.0-RAG system significantly improved the accuracy of DS-related information retrieval. Our evaluations showed that the system achieved over 95% accuracy in answering True/False and multiple-choice questions, outperforming standalone LLMs. Additionally, the user-friendly interface enabled efficient interaction, allowing users to input free-form text queries and receive accurate, contextually relevant responses. The integration process minimized data noise and ensured the most up-to-date and comprehensive DS information was available to users.

**Conclusion:** The integration of iDISK2.0 with an RAG system effectively addresses the limitations of LLMs, providing a robust solution for accurate DS information retrieval. This study underscores the importance of combining structured knowledge graphs with advanced language models to enhance the precision and reliability of information retrieval systems, ultimately supporting better-informed decisions in DS-related research and healthcare.

## Introduction

Dietary supplements (DS) are products intended to add nutritional value to the diet, often containing vitamins, minerals, herbs, amino acids, or other substances^1,2^. The significance of DS has grown considerably in recent years, with a vast majority of individuals recognizing their essential role. According to a survey, approximately nine out of ten dietary or nutritional supplement users agree on the necessity of DS, indicating its widespread acceptance and utilization^3^. DS not only helps in preventing chronic conditions but also holds the potential for significant healthcare savings, as evidenced by research underscoring their economic benefits^4^. Unlike prescription and over-the-counter drugs, DS are primarily considered foods and are regulated by the FDA under different, less stringent rules. The use of DS is often spontaneous rather than based on clinician recommendations, which presents unique challenges in terms of efficacy, safety, regulatory policy, and clinical practice^5^. For instance, approximately 23,000 doctor visits occur annually due to DS-related adverse events^6^. These challenges underscore the need for both consumers and healthcare providers to have access to comprehensive resources that provide reliable DS-related information.

As a new paradigm for managing large-scale heterogeneous biomedical knowledge, biomedical knowledge graphs (BKGs) have been developed using diverse approaches in the past decade^7–9^. In 2020, the International Dietary Supplement Knowledgebase (iDISK) was established to address the growing need for reliable DS information^10^. The iDISK was developed to standardize and integrate dietary supplement information from four existing resources (Natural Medicines Comprehensive Database (NMCD)^11^, “About Herbs” page on the Memorial Sloan Kettering Cancer Center (MSKCC) website^12^, Dietary Supplement Label Database (DSLD)^13^ and Licensed Natural Health Products Database (LNHPD)^14^). The iDISK includes 4208 ingredient concepts linked to drugs, diseases, symptoms, therapeutic classes, organ systems, and products, and it aims to enhance the dissemination of knowledge of dietary supplementation.

However, continual updates in DS-related knowledge, such as changes in product labels and newly discovered interactions between DS ingredients and drugs, have rendered some information in the initial version of iDISK outdated. To ensure the comprehensiveness and accuracy of iDISK, it is essential to upgrade it using the latest versions of source databases and refine integration processes to minimize data noise. With the advent and popularity of large language models (LLMs), many users now turn to these models for DS-related inquiries. Despite their widespread use, LLMs still face challenges in providing accurate knowledge feedback, often resulting in “hallucinations” or incorrect information^15–18^. Retrieval-augmented generation (RAG) combines the robust retrieval capabilities of knowledge bases with the generative prowess of LLMs to enhance the accuracy of responses^19^. A RAG system operates in two primary stages: retrieval and generation^20,21^. The system first retrieves relevant documents or data from a structured knowledge base or database. This ensures that the information being used is accurate, up-to-date, and contextually relevant. The retrieval component often uses advanced algorithms to match user queries with the most pertinent information available in the knowledge base. After retrieving the relevant information, the system uses a generative model (such as an LLM) to produce coherent and contextually appropriate responses. Integrating a KG with an RAG system can provide precise, contextual information while mitigating the hallucination issue inherent in standalone LLMs^22,23^.

Our contributions of this paper are:

1. We presented a significant update from iDISK to iDISK2.0, incorporating the latest database versions and optimizing integration processes to minimize data noise.
2. Integration of an RAG system with LLMs to substantially improve the accuracy of DS question answering task. To the best of our knowledge, this is the first work to introduce RAG and LLM into DS information queries. By combining the generative power of LLMs with precise, context-aware retrieval from a biomedical knowledge graph, we mitigate the risk of misinformation.
3. The development of the iDISK2.0-RAG web portal represents a major advancement, offering both consumers and healthcare providers a reliable, up-to-date resource for DS information query. The portal’s user-friendly interface allows for seamless interaction through simple text inputs, delivering comprehensive and accurate responses in a conversational format. Such framework could further expand to other domains.

Our work with the integration of LLM, RAG, and user interface not only addresses the inherent limitations of standalone LLMs but also sets a new standard for precision and reliability in DS question answering, underscoring the study’s contribution to enhancing the quality and accessibility of DS knowledge. Although this work focuses on DS, our successful integration and platform can provide new opportunities for other domains.

## Methods

Fig.1 illustrates the overall workflow of this study. We first constructed iDISK2.0 by integrating the latest versions of data from four DS resource databases that were originally integrated in iDISK, including NHP, DSLD, MSKCC, and NMCD (Fig 1A). We further developed the Retrieval-Augmented Generation (RAG) based LLM on iDISK2.0 (Fig 1B). In addition, we developed a user-friendly intelligent user interface with iDISK2.0-RAG as the backend to answer DS-related questions (Fig 1C).

**Fig. 1.**
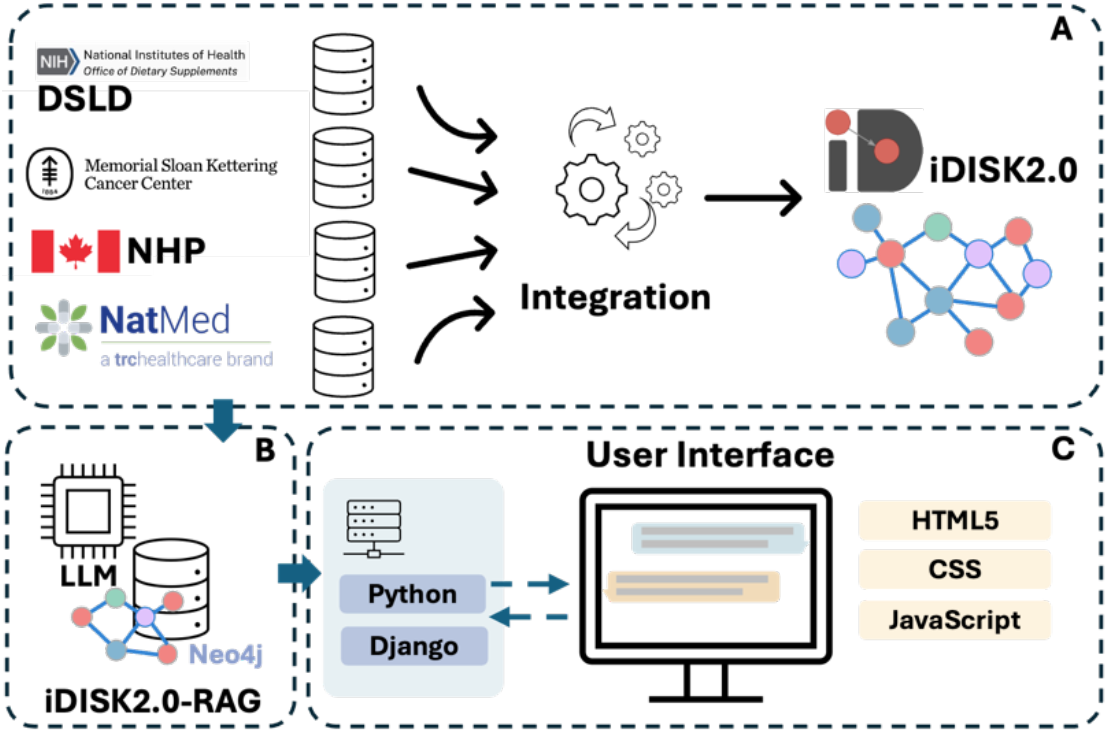
An illustration of the study pipeline

### Data collection and preprocessing

Four datasets were collected and preprocessed for the iDISK 2.0. Data from DSLD was downloaded directly from the data release, and the DSLD API was used to enrich the dietary supplement-related information. Data from MSKCC was obtained after approval, using a web crawler to extract relevant information. LNHPD data was downloaded directly from the data release. After acquiring information from all sources, we converted them into a unified structured format. The data of the NMCD used in the iDISK 2.0 is derived from information that already exists within iDISK. During the information extraction process, we utilized LLM (GPT-4) with the prompt (refer to the supplementary file for specific prompts) to assist in extracting and optimizing information. For example, relationships between DS and diseases recorded in MSKCC were expressed in text format; we used the LLM to extract disease and drug entities from these sentences and organize them into a structured format for integration and processing. The source data contained inconsistencies and noise, such as variations in how company addresses were recorded in the DSLD product data (e.g., “U.S.A.,” “United States of America,” and “United States”) and nonsensical ingredient names like “8” or “%.” We addressed these issues by using LLM to identify and unify the differing expressions. Additionally, we developed filtering patterns specifically designed to remove nonsensical ingredient names, such as those with fewer than two characters, or those consisting solely of numbers or punctuation. This approach ensured a more consistent and accurate dataset by standardizing key information and eliminating irrelevant entries.

Specifically, we extracted the DS product (DSP) name, purpose, safety information, and company address information from LNHPD, along with the DS ingredient (DSI) name, source material information, and relationships between DSP and DSI. From DSLD, we extracted the DSP name, company address information, DSI name, and the relationship between DSP and DSI. In DSLD, names of DSIs appeared as duplicate records due to synonyms (e.g., “Fiber gum acacia,” “Acacia gum extract,” and “Acacia”). We used the DSLD API to collate these synonyms and eliminated duplicates by grouping the synonyms of DSIs representing the same entity using the Ingredient Group classification in DSLD. From MSKCC, we extracted the DSI name, common name, background information, mechanism of action, disease name, drug name, symptom name, and the relationships between DSI and disease, DSI and symptom, and DSI and drug.

After completing the information extraction and optimization, we used QuickUMLS^24^ to map DSI entities, symptom entities, disease entities, and drug entities from the source resources to the UMLS^25^ Concept Unique Identifier (CUI). To ensure mapping accuracy, we restricted each entity type based on UMLS semantic types (refer to the supplementary file). The results of the QuickUMLS mapping included the matched UMLS CUI, term name, similarity, and whether the term was preferred. When QuickUMLS returned multiple results, we employed a prioritization strategy: first, results with a similarity of 1 marked as preferred; next, results with a similarity of 1 even if not marked as preferred; finally, those marked as preferred with the highest similarity. If none of these conditions were met, a UMLS CUI was not assigned to the entity to ensure mapping accuracy.

### Data integration for IDISK2.0

We employed a greedy strategy to standardize entity terminology and integrate data from different sources. For each specific entity type, we initially selected one database to initialize the entity vocabulary. Each specific entity was linked with a unique identifier to integrate entities from all databases, progressively refining the entity vocabulary. Specifically, for DSP, we started with data from LNHPD, integrating DSP entities from both LNHPD and DSLD. We consolidated DSP with the same product name and company name into a single unique entity. We unified entities with the same UMLS CUI or identical term names for DSI. Similarly, we consolidated entities with the same UMLS CUI or identical term names for Disease, Symptom, and Drug entities. Each integrated entity was assigned a unique iDISK ID as its identifier in iDISK. Following this normalization procedure, we obtained a CSV file for each entity type, storing all standardized entity terms and their associated attributes. This allowed us to integrate the extracted knowledge from different databases to build iDISK2.0.

In the knowledge graph (KG), the basic knowledge unit is a triple, typically defined as <head entity, relation, tail entity>, indicating a relationship from the head entity to the tail entity in the KG. We mapped the head and tail nodes of the knowledge from the source databases to iDISK IDs, establishing triples composed of iDISK IDs and their relations. We then deduplicated all these triples to reduce noise in iDISK while ensuring quality. Finally, multiple rounds of manual quality checks were conducted.

### iDISK2.0 deployment with Neo4j

We deployed the upgraded iDISK (iDISK2.0) using Neo4j (https://neo4j.com), a well-designed graph database platform that allows for structured queries within graphs. Specifically, we stored the generated entities and relationships of iDISK in corresponding CSV files, which Neo4j used as input to create a KG instance automatically. This setup enables iDISK2.0 to be efficiently and flexibly interacted with and updated.

### iDISK-based Retrieval-Augmented Generation (RAG) system

Fig. 2 illustrates the overall framework of iDISK-RAG. We began by preprocessing all entities within the iDISK2.0 database, creating embedding vectors for each entity name using OpenAI’s embedding model, text-embedding-3-small (Fig. 2A). This step allowed us to construct the iDISK2.0 entity vector database, which serves as a foundation for efficient and accurate entity matching. When a user inputs a query, we first extract relevant entities from the question by employing an LLM (GPT-4.0) (Fig. 2B), using tailored prompts as follows:

> - *You are an expert entity extractor from a sentence in the biomedical domain. Please identify the entity from the provided sentence and return that entity name. The entity can only be the following types: Dietary Supplement Ingredient, Drug, Disease, Symptom, Therapeutic Class, System Organ Class, and Dietary Supplement Product. Also, please add your identified entity type after the entity name with “:”. For example, if the provided sentence is: “Out of the given list, which disease is Caffeine effective?” Your response should be: Caffeine: Dietary Supplement Ingredient. If you recognize more than one entity from the sentence, please use “* || *“ to split those identified entities. For example, if the provided sentence is: “Out of the given list, what is the relationship between Coenzyme Q10 and Ischemia-reperfusion injury?” Your response should be: Coenzyme Q10: Dietary Supplement Ingredient* || *Ischemia-reperfusion injury: Disease*.

**Fig. 2.**
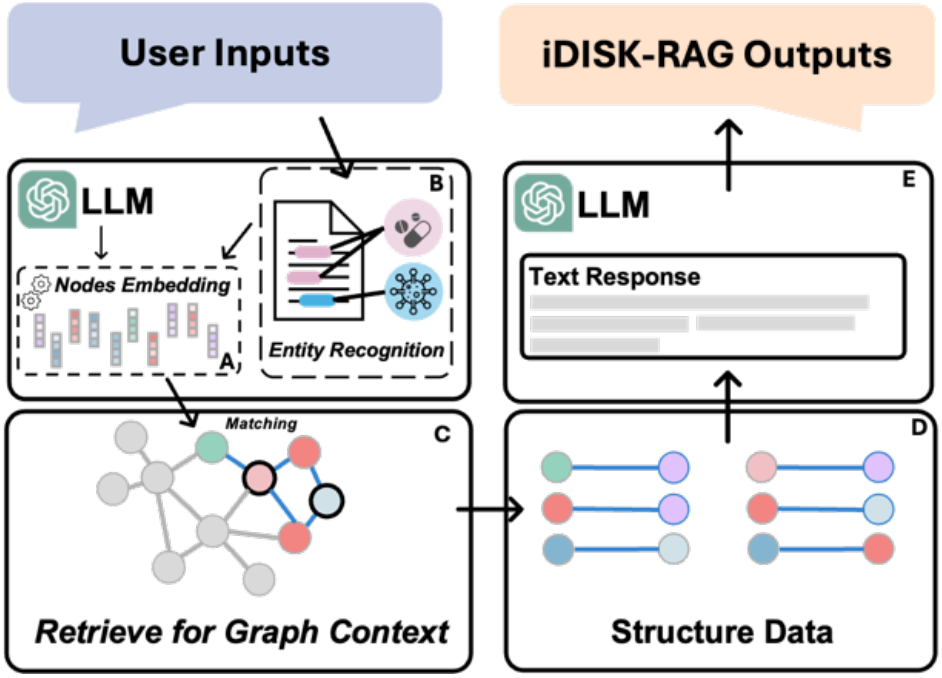
The overall design of the iDISK-RAG

The entities identified through this process are then transformed into corresponding vectors using the same OpenAI embeddings model.

To match these identified entities with those in the iDISK2.0 database, we calculate the cosine similarity between the vectors of the identified entities and the vectors in the preprocessed iDISK2.0 entity vector database. Cosine similarity, a measure of similarity between two vectors in an inner product space, is calculated based on the cosine of the angle between them (detailed information on cosine similarity is provided in the supplementary file). We then select the entity with the highest similarity score, provided that the score is 0.75 or higher, to ensure a reliable match between the identified entity and a corresponding entity in iDISK2.0.

Once a match is identified, the corresponding nodes are converted into Cypher queries, allowing for interaction with iDISK2.0, which is deployed on the Neo4j graph database platform. This process retrieves the relevant graph context (Fig. 2C), which is returned as structured data in the form of triples (Fig. 2D). Finally, an LLM is used to integrate the user’s original question with the retrieved structured data (triples) and generate a comprehensive and contextually enhanced response (Fig. 2E). The prompt used for answer generation is:

> - *You are an expert biomedical researcher. To answer the question at the end, you need to first read the provided structured knowledge. The format of the structured knowledge is: (entity)-[relationship]-(entity). You should understand the provided structured knowledge and choose one or more knowledge as the basis to answer the corresponding question. For example, the question is: Which disease is Butterbur effective? And the provided structured knowledge is: [‘(Butterbur)-[is_effective_for]-(Hay fever)’, ‘(Butterbur)-[is_effective_for]-(Migraines)’]. Your answer should be: Migraines. If the structural knowledge provided is empty, please first give the following feedback: Failed to retrieve relevant knowledge in iDISK2*.*0. Then answer the question based on your own knowledge. If the question is: Which disease is L-Arginine effective? And the provided structured knowledge is: [‘(L-Arginine)-[is_effective_for]-(Gestational Hypertension),’ ‘(L-Arginine)-[is_effective_for]-(Hypertension),’ ‘(L-Arginine)-[is_effective_for]-(Erectile dysfunction),’ ‘(L-Arginine)-[is_effective_for]-(Necrotizing enterocolitis),’ ‘(L-Arginine)-[is_effective_for]-(Pre-eclampsia),’ ‘(L-Arginine)-[is_effective_for]-(Peripheral arterial disease)’]. Your answer should be: Failed to retrieve relevant knowledge in iDISK2*.*0. However, based on my knowledge that … For example, the question is: Is it true that Vitamin D is effective for Osteoporosis? And the provided structured knowledge is: [‘(Vitamin D)-[is_effective_for]-(Osteoporosis)’]. Your answer should be: True, If the question is: Is it true that Flaxseed is effective for Wounds? And the provided structured knowledge is: []. Your answer should be: Failed to retrieve relevant knowledge in iDISK2*.*0. However, based on my knowledge that* …

### Evaluation

We extracted two types of questions from information on the MSKCC website: True/False (T/F) and Multiple-Choice Questions (MCQ). Specifically, we extracted information regarding the effectiveness of DS and diseases and the interactions between DS and drugs. This information was formatted into T/F and MCQ questions (473 T/F questions and 329 MCQs). To evaluate the performance of iDISK-RAG and two LLMs (GPT-3.5 and GPT-4.0), we randomly selected 100 questions from each type (T/F and MCQ). These questions were answered by iDISK-RAG, GPT-3.5, and GPT-4.0, and their accuracy was calculated based on the correct answers. This process was repeated ten times using bootstrapping, resulting in each system answering 1000 T/F questions and 1000 MCQ questions. Accuracy metrics for each sampling iteration were then computed to derive the performance distribution. To ensure a fair evaluation, all knowledge from MSKCC was excluded from the retrieval stage in the RAG framework, preventing any influence of MSKCC-integrated knowledge on the test results. The rose plot was drawn using ChiPlot^26^.

### RAG-enhanced intelligent visual interfaces

We developed a web-based graphical portal that allows users to access knowledge from iDISK2.0 intuitively and flexibly. Specifically, we built the backend (server-side) using Django (https://www.djangoproject.com/), an advanced web framework based on Python. The backend also hosts our iDISK-RAG framework to provide users with more accurate retrieval information. The front end (web application side) is constructed using HTML5 and CSS, with JavaScript employed for data interaction. Specifically, when the backend receives a query from the front end, the system first utilizes an LLM, such as GPT-4.0, to extract relevant entities from the user’s question through tailored prompts. These extracted entities are then matched with pre-trained embedding information from a comprehensive entity vocabulary stored in the backend. Once a match is identified, the RAG system generates corresponding Cypher queries based on the matched entities. These Cypher queries are subsequently used to interact with iDISK2.0, which is deployed on the Neo4j graph database platform, to retrieve the relevant contextual subgraphs. The retrieved subgraphs provide structured data that is essential for generating an accurate response. In the final step, the LLM, guided by specific prompts, is employed once again to transform the structured data retrieved by the RAG system into natural language, which is then returned to the front end’s dialogue interface. In cases where the entities mentioned in the user’s question cannot be successfully matched within iDISK2.0, the LLM (GPT-4.0) relies on its internal knowledge base to generate a corresponding answer, which is also delivered to the front end’s dialogue interface. This approach ensures that users receive comprehensive and contextually relevant responses, whether through structured data retrieval or the LLM’s generative capabilities.

## Result

### The integrated DIetary Supplements Knowledge Base 2.0

Through systematic collection and coordination, we ultimately obtained 279 concepts of DSIs, 270 concepts of Drugs, 231 concepts of Diseases, and 425 concepts of Symptoms from the latest version of the MSKCC website. From the DSLD database, we acquired 4,318 concepts of DSIs and 92,651 concepts of DSPs. Additionally, from the NHP database, we gathered 4,690 concepts of DSIs and 71,364 concepts of DSPs. After conducting data management and data cleaning, we integrated data from the three sources through biomedical entity term normalization and knowledge integration. The current version of iDISK encompasses 174,317 entities across seven types (Table 1), including 8,091 DSI entities, 163,806 DSP entities, 786 disease entities, 625 drug entities, 425 symptom entities, 567 dietary therapeutic class (TC) entities, and 17 System Organ Class (SOC) entities. Additionally, there are six types of relationships among the seven entity types (Table 1), including DSP-DSI, DSI-disease, DSI-symptom, DSI-drug, DSI-TC, and DSI-SOC.

**Table 1.** The concept, relationship, and attribute in iDISK2.0.

| Concept | iDISK (old version) | iDISK (new version) |
| --- | --- | --- |
| Dietary Supplement Ingredient | 4,208 | 8,091 |
| Dietary Supplement Product | 137,568 | 163,806 |
| Drug | 495 | 625 |
| Disease | 776 | 786 |
| Therapeutic Class | 605 | 567 |
| System Organ Class | 17 | 17 |
| Sign / Symptoms | 985 | 425 |
| **Total** | **144,654** | **174,317** |
| **Relationship** |  |  |
| is effective for | 5,363 | 5,245 |
| has therapeutic class | 5,454 | 4,435 |
| has adverse effect on | 3,168 | 2,598 |
| has adverse reaction | 2,233 | 1,342 |
| has ingredient | 689,826 | 317,062 |
| interacts with | 3,631 | 3,583 |
| **Total** | **709,675** | **334,265** |
| **Attribute** |  |  |
| Company Name | - | 163,806 |
| Company Address | - | 163,806 |
| Product Purpose | - | 65,097 |
| Product Risk | - | 63,623 |
| Background | 1,399 | 1,289 |
| Safety | 1,219 | 1,179 |
| Mechanism of action | 258 | 277 |
| Source Material | 5,532 | 4,277 |
| Interaction Rating | 3,076 | 3,086 |
| Effectiveness Rating | 4,307 | 4,623 |
| **Total** | **15,791** | **471,063** |

The current version of iDISK also includes 471,063 attributes (Table 1), comprising 163,806 DSP company names, 163,806 DSP company addresses, 65,097 DSP purposes, 63,623 DSP risks, 1,289 DSI backgrounds, 1,179 DSI safety profiles, 277 DSI mechanisms of action, 4,277 DSI source materials, 3,086 DSI-drug interaction ratings, and 4,623 DSI-disease effectiveness ratings.

We deployed iDISK2.0 using Neo4j. The entity and relationship source files of iDISK2.0 have been published in comma-separated values (CSV) format and are available at the following URL: https://github.com/houyurain/iDISK2.0. It is important to note that the deployed version of iDISK2.0 excludes data from NMCD due to certain restrictions.

### A iDISK2.0-based Retrieval-Augmented Generation (RAG) system

To address the potential hallucination issue present in current LLMs and thereby improve the accuracy of user responses, we developed an iDISK-based RAG system. This system integrates the strengths of KG and LLMs to efficiently retrieve relevant knowledge from iDISK and utilize the language model to generate feedback and facilitate user interaction.

We collected data from the MSKCC website regarding the effectiveness of dietary supplements and diseases and the interactions between dietary supplements and drugs. From this data, we generated a total of 473 T/F questions and 329 MCQs. Specifically, there are 164 T/F questions and 123 MCQs related to dietary supplements and diseases, and 309 T/F questions and 206 MCQs related to dietary supplements and drugs. For instance, in the MSKCC database, there is a record stating, “Vitamin C is used to prevent and treat the common cold.” We utilized this record to generate corresponding True/False and MCQs for evaluative purposes. The True/False question derived from this statement is: “*Is it true that Vitamin C is effective for the common cold*?” with the correct answer being

“*True*.” Additionally, we developed MCQ to assess understanding: “O*ut of the given list, which disease is Vitamin C effective for?*” The provided options are: *Bladder stones, Common cold, Stroke, Bleeding hemorrhoids, None of the above*. The correct answer is “*Common cold*.” Fig. 3 illustrates the performance (accuracy) of two LLMs (GPT-3.5 and GPT-4.0) and the iDISK-RAG system on T/F and MCQs related to dietary supplements and diseases, as well as dietary supplements and drugs. The results show that iDISK-RAG achieves over 95% accuracy across all question types: 99% for T/F questions on dietary supplements and diseases, 99% for MCQs on dietary supplements and diseases, 97% for T/F questions on dietary supplements and drugs, 95% for MCQs on dietary supplements and drugs. In contrast, the performance of the two LLMs is notably lower, with GPT-4.0 outperforming GPT-3.5: GPT-4.0 achieved 93% accuracy on T/F questions related to dietary supplements and diseases, compared to 85% for GPT-3.5, 73% accuracy on MCQs related to dietary supplements and diseases, compared to 45% for GPT-3.5, 62% accuracy on T/F questions related to dietary supplements and drugs, compared to 40% for GPT-3.5, 52% accuracy on MCQs related to dietary supplements and drugs, compared to 46% for GPT-3.5. Despite GPT-4.0’s superior performance over GPT-3.5, both models fall significantly short of the accuracy achieved by iDISK-RAG in these specialized domains.

**Fig. 3.**
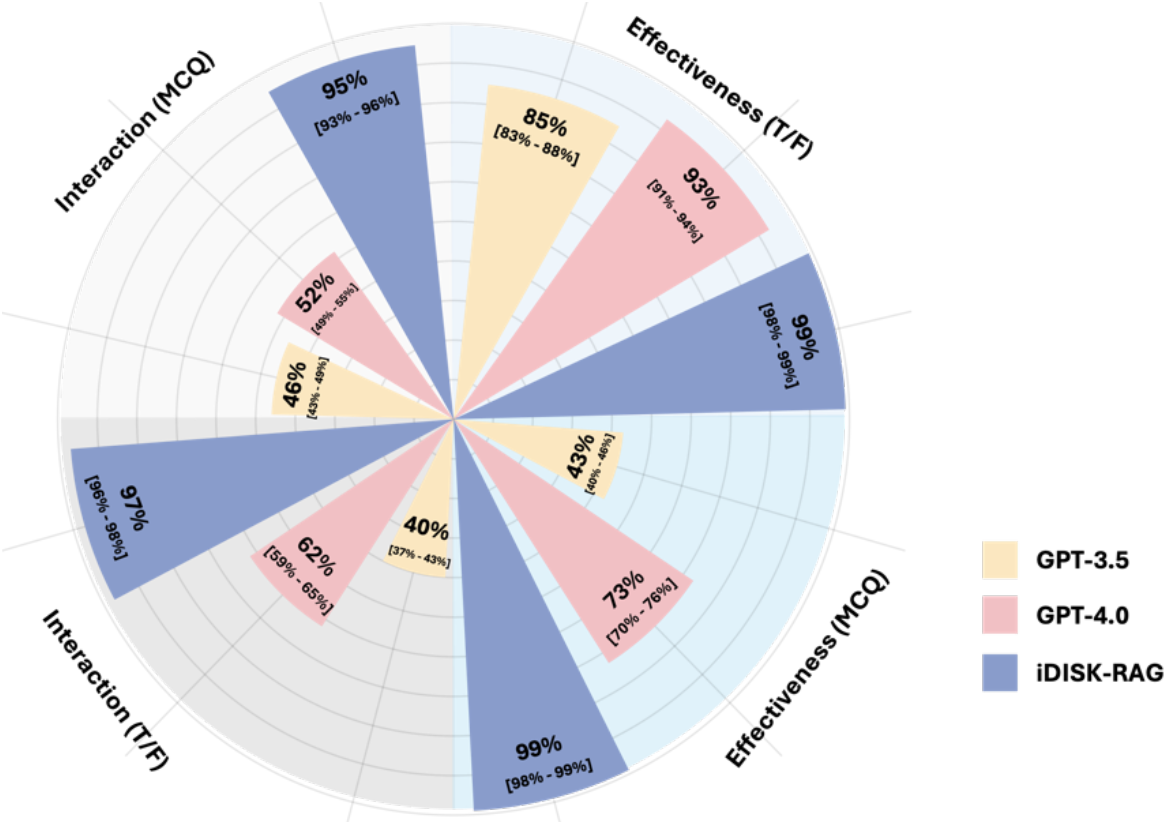
Model performance in Q-A tasks

### A user-friendly intelligent user interface for knowledge retrieval

Knowledge retrieval is one of the most common application scenarios of BKG in biomedical research. In recent years, LLMs have shown outstanding performance in NLP tasks but suffer from the issue of “hallucinations,” especially within specific domains^15–17^. To facilitate the use of iDISK2.0 by biomedical and clinical researchers in DS studies, we developed a web portal based on iDISK2.0-RAG as the backend. This RAG-based UI allows users to input free-form text queries, generating meaningful biomedical text responses based on the established knowledge within iDISK2.0 to answer users’ questions. Figure 4A illustrates an example of a user query using free-form text. When the user inputs a question such as “Which disease is Omega-3 Fatty Acids effective for?”, the UI retrieves relevant knowledge from the iDISK2.0-RAG backend and returns the corresponding answer: “Omega-3 Fatty Acids are effective for: -Cardiovascular disease - Lupus - Cancer - Depression – High cholesterol - Atherosclerosis.” In cases where the user’s query cannot be answered directly from the iDISK2.0 knowledge base, the RAG framework informs the user that the specific knowledge is not available within iDISK2.0. Subsequently, the LLM within the framework attempts to answer the question based on its own knowledge, as depicted in Figure 4B.

**Fig. 4.**
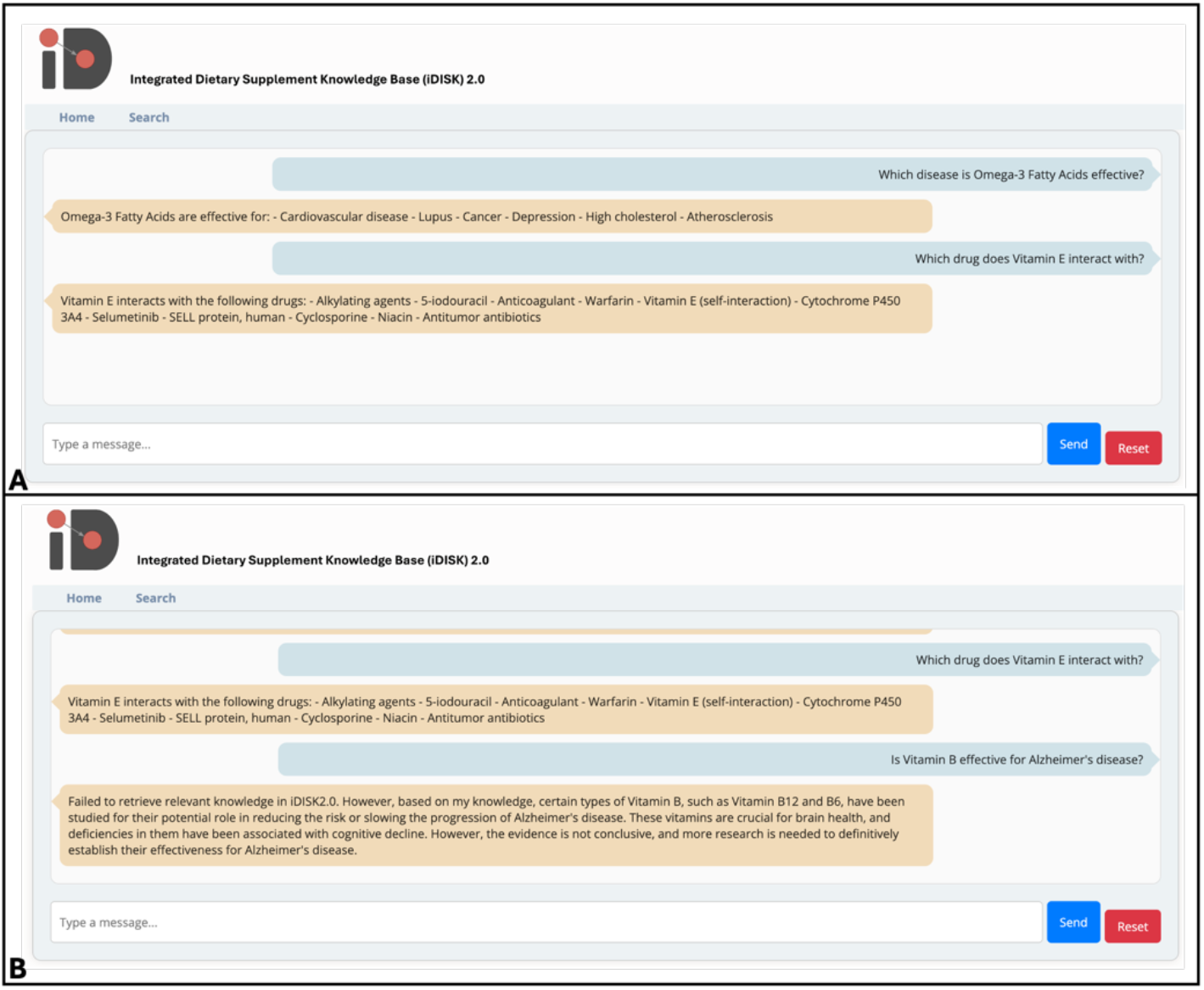
An example of Q-A on UI

## Discussion

iDISK integrates information from four well-designed DS databases, ensuring it contains the most comprehensive DS-related information available. iDISK2.0 inherits this feature, maintaining its status as the most thorough DS information repository. As source resources are updated, much DS-related information has been upgraded. With ongoing research, DS information continues to iterate and expand. For instance, when the DSLD was first launched in 2013, it contained nearly 17,000 label entries; now it records nearly 190,000, an increase of more than tenfold in just a decade. Additionally, many DS details have been corrected; for example, the older version of iDISK included an ingredient from DSLD labeled as “Araceae,” which was inaccurate. In the latest version of DSLD, this ingredient has been removed. iDISK2.0 ensures users have access to not only the most comprehensive but also the most up-to-date DS information. Upgrading iDISK to iDISK2.0 is crucial to maintaining the accuracy and relevance of DS information. Without regular updates, the knowledge base could become outdated, leading to potential misinformation and inefficiencies in DS-related research and healthcare decisions. The continuous integration of the latest data and refined processes in iDISK2.0 helps mitigate these risks, ensuring that users have reliable, current information to make informed decisions about dietary supplements.

LLMs like GPT-4.0 have significantly advanced the field of natural language processing. However, they still exhibit notable limitations, particularly in domain-specific applications such as DS information retrieval. A key challenge is their propensity for generating “hallucinations”—plausible yet incorrect information—stemming from their reliance on extensive, but often unverified, data sources^18^. This issue is particularly problematic when precision is critical. For example, our evaluation highlighted a case involving the use of Coenzyme Q10, also known as Ubiquinone, which is documented for its effectiveness in preventing migraines according to the MSKCC database. To test our system’s accuracy, we posed a multiple-choice question: “Which disease is Ubiquinone effective for?” with options including Migraines, Hypertension, Menstrual disorders, Cholestasis, and None of the above. Despite the correct answer being “Migraines,” based on robust evidence. However, GPT-4.0 returned “Cholestasis” as the answer, even though only limited studies have explored the relationship between Coenzyme Q10 and Intrahepatic Cholestasis of Pregnancy (ICP)^27,28^, with no definitive conclusions indicating its efficacy in this context. In contrast, numerous studies have established the effectiveness of Coenzyme Q10 in the prevention and treatment of migraines^29–31^. This error underscores a critical concern: while LLMs have vast knowledge bases, they can still falter by either misinterpreting context or lacking explicit, well-established information.

To address these challenges, we developed iDISK2.0 and integrated it with a RAG system. This integration combines the expansive knowledge base of LLMs with the precise, validated data from iDISK2.0, significantly reducing the risk of hallucinations. In testing, the RAG system, drawing from verified knowledge within iDISK2.0, consistently provided accurate answers, such as correctly identifying Coenzyme Q10’s efficacy for migraines. Further enhancing this solution, we incorporated a user-friendly interface with the iDISK2.0-RAG backend, allowing users to access accurate DS information through simple conversational text inputs. This setup not only facilitates better-informed decisions but also improves the overall user experience by ensuring the dissemination of the most accurate and current DS information. The necessity of this innovation is emphasized by the potential risks of relying solely on LLMs, which can propagate misinformation and lead to inefficiencies in both research and healthcare decisions. Moving forward, future research should focus on refining the interaction between LLMs and RAG systems. One potential avenue could involve mechanisms that allow LLMs to defer to the more reliable RAG output in cases where the evidence is more compelling. This approach would significantly enhance the accuracy and trustworthiness of AI-generated responses in biomedical applications.

Despite adopting stricter mapping and integration schemes compared to the older version of iDISK, some errors and noise still exist in the final results. These inaccuracies partly stem from the imprecise expressions in the source databases. Additionally, errors arise when attempting to map entities from the source databases to UMLS CUIs for subsequent entity integration. The entity extraction and mapping software QuickUMLS sometimes returns incorrect results. For instance, the MSKCC database records an interaction between “Andrographis” and “Blood pressure-lowering drugs”: “Andrographis interacts with Blood pressure-lowering drugs.” When extracting entities from this knowledge, we extracted the drug entity “Blood pressure-lowering drugs” from the text. However, QuickUMLS mapped “Blood pressure-lowering drugs” to “Drug (UMLS CUI: C0013227),” resulting in an incorrect mapping and introducing errors into the integration process.

Given the current limitations of this study, we plan to implement different strategies in the future to further reduce errors. For instance, we can introduce entity extraction techniques based on LLMs to extract DS-related information stored in text. For example, we may employ various prompt strategies, including rule-based prompts (e.g., HealthPrompt^32^), hard-prompt learning^33^ and soft-prompt learning^34^. Additionally, we can leverage LLMs to batch-check entity extraction and mapping results and perform quality control on the integration outcomes. Recent research has highlighted the beneficial applications of BKGs in the medical field^7,35^. Moreover, the rise of LLMs has spurred discussions on how best to combine the strengths of these models and BKGs^36^. In the future, besides continuously maintaining iDISK to ensure it remains the most up-to-date and comprehensive DS information resource, we will also explore ways to better assist users in utilizing iDISK. For example, we plan to introduce knowledge graph embedding (KGE) technology. By using graph-structured reasoning, we aim to offer reasonable predictions for knowledge not yet included in existing databases, thereby helping users discover more information. This approach will not only enhance the precision of user queries but also expand the scope of knowledge available to them.

## Conclusion

In this study, we developed iDISK2.0, a comprehensive and up-to-date repository for DS information, by integrating the latest data from NMCD, MSKCC, DSLD, and LNHPD. We addressed the limitations of LLMs in accurately answering DS-related queries by creating an RAG system that leverages both LLMs and a BKG. Our evaluation demonstrated that iDISK2.0-RAG outperforms standalone LLMs, achieving over 95% accuracy in various DS-related questions. While some errors and noise persist due to imprecise source data and entity mapping challenges, future improvements will focus on advanced entity extraction and quality control techniques. We aim to continuously maintain and enhance iDISK2.0, supporting users in making informed DS-related decisions through precise and expanded knowledge.

## Data Availability

All data produced are available online at

https://github.com/houyurain/iDISK2.0

## Author Contributions

YH and RZ conceived the study design and wrote the manuscript. YH implemented the experiments of the study.

